# Within-host modelling of primaquine-induced haemolysis in hemizygote glucose-6-phosphate dehydrogenase deficient healthy volunteers

**DOI:** 10.1101/2024.08.01.24311380

**Authors:** James A Watson, Parinaz Mehdipour, Robert Moss, Podjanee Jittamala, Sophie Zaloumis, David J Price, Saber Dini, Borimas Hanboonkunupakarn, Pawanrat Leungsinsiri, Kittiyod Poovorawan, Kesinee Chotivanich, Germana Bancone, Robert J Commons, Nicholas PJ Day, Sasithon Pukrittayakamee, Walter RJ Taylor, Nicholas J White, Julie A Simpson

## Abstract

Primaquine is the only widely available drug to prevent relapses of *Plasmodium vivax* malaria. Primaquine is underused because of concerns over oxidant haemolysis in glucose-6-phosphate dehydrogenase (G6PD) deficiency. A pharmacometric trial showed that ascending-dose radical cure primaquine regimens causing ‘slow burn’ haemolysis were safe in G6PD deficient male volunteers. We developed and calibrated a within-host model of primaquine haemolysis in G6PD deficiency, using detailed serial haemoglobin and reticulocyte count data from 23 hemizygote deficient volunteers given ascending-dose primaquine (1,523 individual measurements over 656 unique timepoints). We estimate that primaquine doses of ∼0.75mg base/kg reduce the circulating lifespan of deficient erythrocytes by ∼30 days in individuals with common Southeast Asian *G6PD* variants. We predict that 5mg/kg primaquine total dose can be administered safely to G6PD deficient individuals over 14 days with expected haemoglobin drops of 18 to 43% (2.7 to 6.5g/dL drop from a baseline of 15g/dL).

## Background

Glucose-6-phosphate dehydrogenase (G6PD) deficiency is the most common enzymopathy of humans [1]. It is prevalent in areas where malaria is, or once was, endemic. G6PD deficiency is the major obstacle to the control and elimination of relapsing vivax malaria. *P. vivax* is now the predominant cause of malaria in much of Asia, the Americas and the horn of Africa [2]. The 8-aminoquinoline drugs, primaquine and tafenoquine, are the only available antimalarial drugs which have anti-relapse activity (radical cure) in vivax malaria, but they cause dose-dependent haemolysis in G6PD deficiency [3]. Because G6PD testing is often not available in routine care, patients are not prescribed 8-aminoquinoline treatment, resulting in substantial morbidity and contributing to mortality [4].

Radical curative efficacy is proportional to the total dose of primaquine administered. The World Health Organisation (WHO) recommends that in patients with *P. vivax* or *P. ovale* malaria who are G6PD deficient (usually defined as *<*30% of the adjusted male median enzyme activity), primaquine should be given over 8 weeks as a weekly dose of 0.75mg base/kg [5]. The rationale for this dosing regimen is based on the pharmacodynamics of 8-aminoquinoline-induced haemolysis. Mutations in the *G6PD* gene causing G6PD deficiency result in an unstable enzyme. Older red blood cells become G6PD depleted and thus more vulnerable to oxidative haemolysis [6, 7]. The 0.75mg/kg (adult dose 45mg) weekly dosing regimen was designed to provide sufficient time for compensatory erythropoeisis which reduces the overall fall in haemoglobin, but at the expense of a protracted (8 week) treatment course. This 8-weekly dosing regimen was chosen on the basis of a small haematological study in three individuals who most likely had *G6PD* African A-variants [8]. Its safety has not been established in patients with more severe variants of G6PD deficiency. Efficacy depends on good adherence over the 8 weeks, but nearly all the haemolytic risk is incurred from the first doses, requiring clinical monitoring [9, 10].

To develop shorter primaquine dosing regimens with acceptable safety profiles for G6PD deficient patients with vivax malaria, we initially compared ascending primaquine dose regimens with a single high dose of primaquine (45mg) in hemizygote G6PD deficient male volunteers in Thailand [11]. Ascending primaquine dose regimens are theoretically the safest way of administering radical curative dose regimens [12]. By inducing ‘slow burn’ haemolysis, they allow time for compensatory erythropoeisis, enabling the older red cell population to be replaced by younger cells with higher intraerythrocytic G6PD activities and increased oxidant resistance. In order to characterise the red cell dynamics following ascending primaquine doses, we developed a within-host Bayesian pharmacodynamic model of red blood cell production and turnover in G6PD deficiency. We fitted this model to the data from the healthy volunteer study and inferred the primaquine dose-dependent reduction in red cell lifespan. This model allows us to make generalisable predictions about the haemolytic response to other ascending dose primaquine regimens, in order to determine optimal dosing strategies for radical cure in patients with relapsing malarias and G6PD deficiency.

## Results

### Overview of study data

The two studies enrolled 27 healthy male Thai and Burmese G6PD deficient volunteers. 24 volunteers received ascending primaquine doses, and 16 volunteers received single 45mg doses, of whom 13 participated in both studies. One participant in the ascending dose study was excluded from this analysis: he only received three 7.5mg base primaquine doses and then was withdrawn from the study due to severe back pain (prolapsed intervertebral disc – unrelated to primaquine). He was subsequently lost to follow-up. Given the very low total dose he received, his data are non-informative. The baseline characteristics of the 26 volunteers included in the analysis are shown in Table 1. Figure 1 shows the haemoglobin and reticulocyte data from the 26 volunteers by study, coloured by either the day 10 cumulative dose in the ascending dose study (a proxy for how quickly the dose was increased); or the mg/kg single dose.

**Table 1:** Baseline characteristics of the healthy male G6PD deficient volunteers. For the continuous variables we show the median (range). Of the 26 volunteers included in the analysis, 13 participated in both sub-studies. ^†^also known as Quing Yan [13].

|  | Part 1 - Ascending dose | Part 2 - Single 45mg dose | Overall |
| --- | --- | --- | --- |
| *n* | 23 | 16 | 26 |
| Age (years) | 32 (18-55) | 34 (20-58) | 32 (18-58) |
| Weight (kg) | 64 (46-86) | 64 (52-86) | 64 (46-86) |
| *G6PD* genotype |  |  |  |
| Viangchan (871G>A) | 12 | 6 | 12 |
| Mahidol (487G>A) | 3 | 2 | 3 |
| Canton (1376G>T) | 4 | 3 | 5 |
| Aures (143T>C) | 1 | 1 | 1 |
| Chinese-4 (392G>T)<sup>†</sup> | 1 | 0 | 1 |
| Orissa (131C>G) | 1 | 1 | 1 |
| Union (1360C>T) | 1 | 2 | 2 |
| Kaiping (1388G>A) | 0 | 1 | 1 |
| *G6PD* enzyme activity (U/g Hb) | 0.15 (0-1.9) |  |  |
| Haemoglobin (g/dL) | 14.3 (11.8-15.8) | 14.0 (12.3-15.9) |  |
| Red cell count (x10<sup>12</sup> per L) | 4.9 (4.2-6.0) | 5.1 (3.9-5.9) |  |
| Reticulocyte count (%) | 2.4 (1.1-4.0) | 2.4 (1.0-2.9) |  |
| *CYP2D6* genotypes |  |  |  |
| *10/*10 | 6 | 4 | 8 |
| *2/*10 | 6 | 4 | 7 |
| *1/*10 | 6 | 4 | 7 |
| *1/*2 | 3 | 3 | 3 |
| *1/*1 | 2 | 1 | 2 |

**Figure 1:**
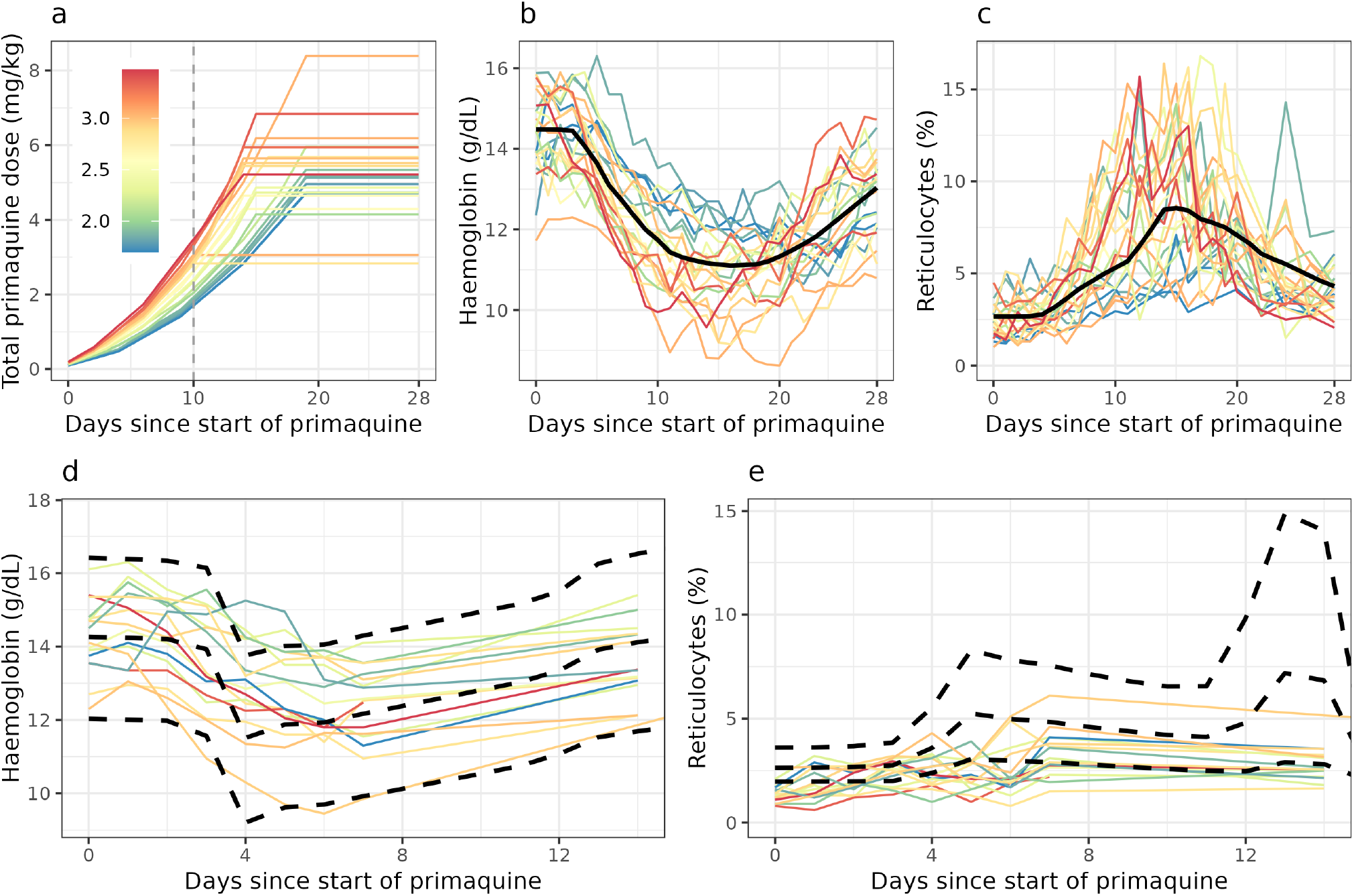
Data from the primaquine challenge study. Top panels show data from Part 1 study (ascending dose regimens); the bottom panels show Part 2 study (single 45mg dose). Colours correspond to the day 10 cumulative dose for the ascending dose study (proxy for how quickly the primaquine dosing was escalated), and to the mg/kg dose for the single dose study. Solid black lines indicate the mean model fit for the ascending dose study (top panels). Dashed black lines in the bottom panels indicate the mean model predictions and 90% predictive intervals for the single dose study, generated from the model fit for the ascending dose study.

### Within host model of primaquine-induced haemolysis

We developed a within-host model of primaquine-induced haemolysis in G6PD deficiency and compensatory erythropoeisis (see Methods for details on the model structure and implementation). Figure 2 shows an overview of the model structure. The key assumptions driving the model behaviour are: (i) the haemolytic effect of primaquine is mediated by a dose-dependent reduction in the lifespan of circulating erythrocytes; (ii) hysteresis in the haemolytic effect of primaquine can be described as a weighted sum of the doses given over the previous 10 days (non-parametric delay in effect); (iii) compensatory erythropoeisis is a function of both the absolute difference from the steady state haemoglobin, and the daily drop in haemoglobin (absolute haemolysis and rate of haemolysis).

**Figure 2:**
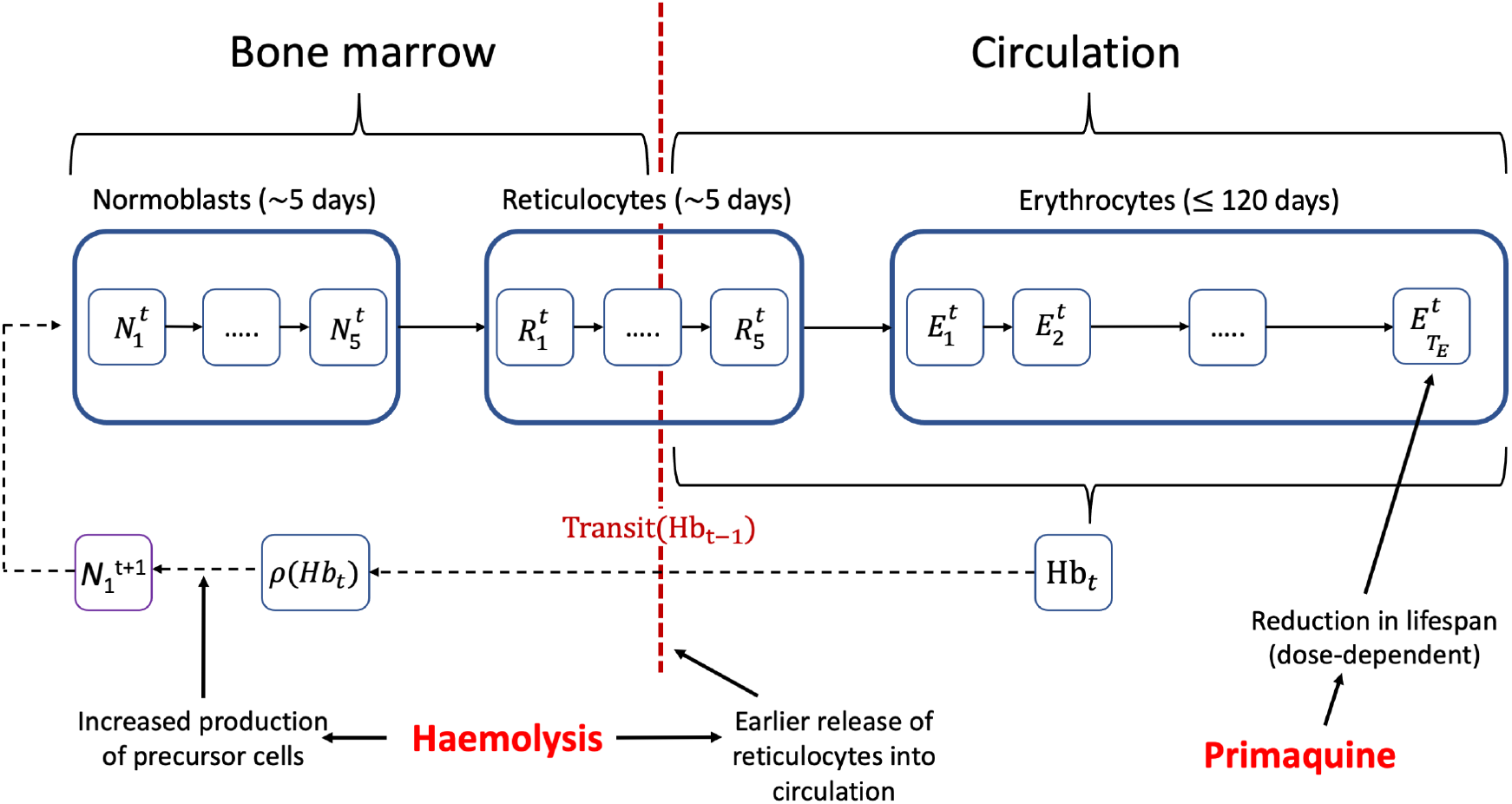
Compartmental model of red blood cell turnover and primaquine-induced haemolysis. Each compartment corresponds to one day and the model iterates daily. Primaquine administration reduces the lifespan of the erythrocytes resulting in haemolysis; haemolysis leads to greater production of precursor cells and earlier release of reticulocytes into circulation.

We fitted the model in a Bayesian framework to the serial haemoglobin and reticulocyte data from the 23 volunteers who were given ascending primaquine regimens. The data included all haemoglobin and reticulocyte measurements taken between day 0 (day of first primaquine dose) and day 28, a total of 656 individual timepoints with 1,523 individual measurements. Estimates of key model parameters are presented in Table 2. Overall, the model captured the individual profiles for both the haemoglobin and the reticulocyte counts well. To check the predictive accuracy of the model, we re-fitted it 23 times, each time leaving one participant out, and then predicted their haemoglobin and reticulocyte profiles based on the primaquine regimen they took. This showed that the model could reliably predict out-of-sample data (Figures 3 and 4). One individual (subject 11) had observed haemolysis which was substantially greater than the model prediction. He was *G6PD* Union (131C>G) with a normal *CYP2D6* genotype (*1*/*\*10). Primaquine was stopped in this participant on day 10 because he met the prespecified stopping rule (40% reduction from baseline haemoglobin).

**Table 2:** Summary of key parameter estimates from the model fitted to the ascending dose study. We report population parameters as the mean (95% credible intervals [CrI]). Inter-individual variation is reported as 80% predictive intervals.

| Parameter | Population mean (95% CrI) | Inter-individual variation (10<sup>th</sup> to 90<sup>th</sup> percentiles) |
| --- | --- | --- |
| Steady-state haemoglobin (g/dL) | 14.5 (14.1-14.9) | 13.3 to 15.7 |
| Steady-state RBC lifespan (days) | 60 (55-65) | 46 to 74 |
| Max RBC lifespan reduction (%) | 36 (31-41) | 25 to 47 |
| Half-maximal effect dose (mg/kg) | 0.18 (0.15-0.22) | 0.12 to 0.28 |
| Dose-response slope | 2.25 (1.81, 2.69) | — |

**Figure 3:**
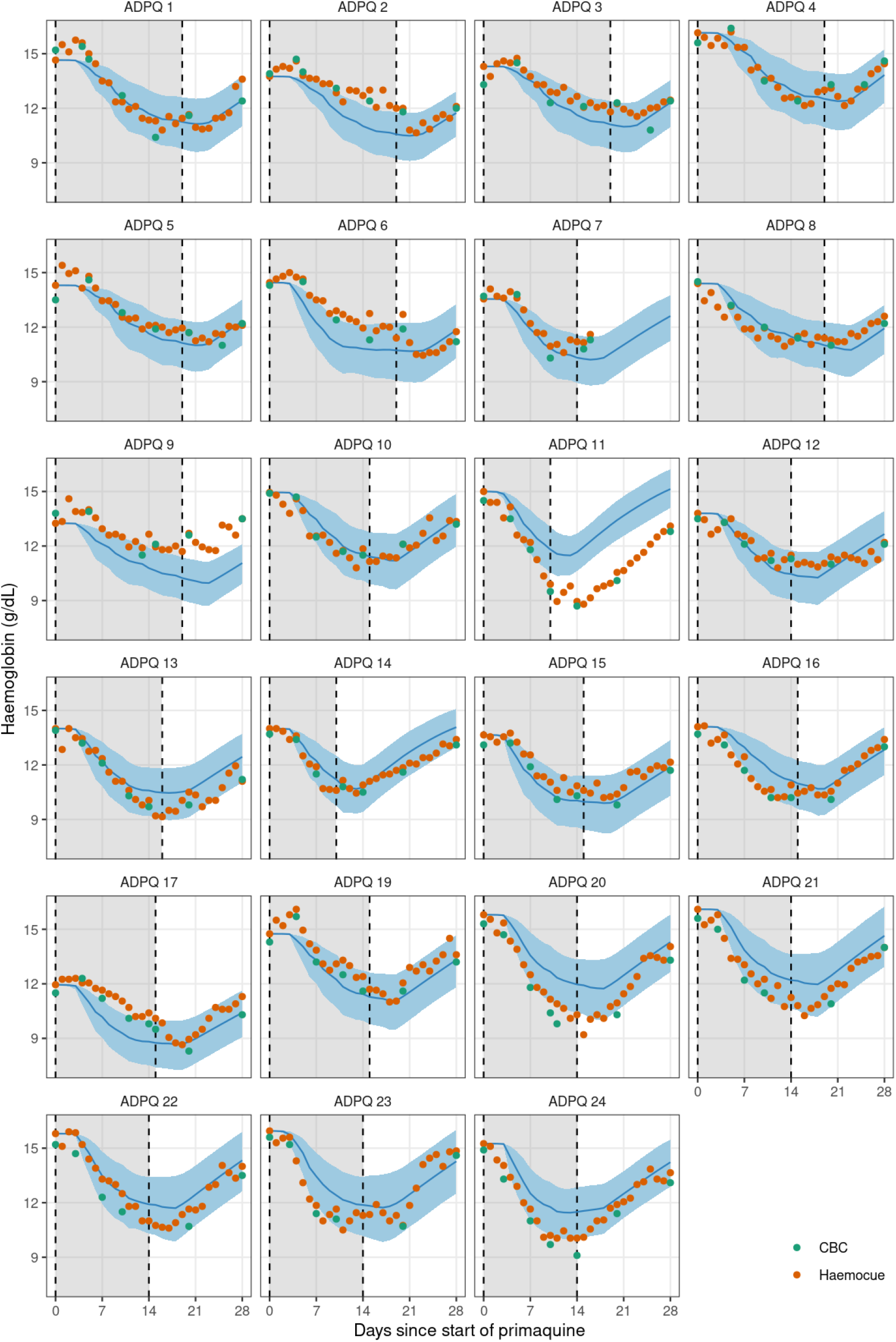
Leave-one-out predictions for the haemoglobin concentrations over time for each individual in the ascending-dose study. Dots show the haemoglobin measurements (green: CBC; orange: haemocue). Blue lines (shaded areas) show the mean model prediction (90% predictive intervals). The grey shaded area indicates the primaquine dosing period.

**Figure 4:**
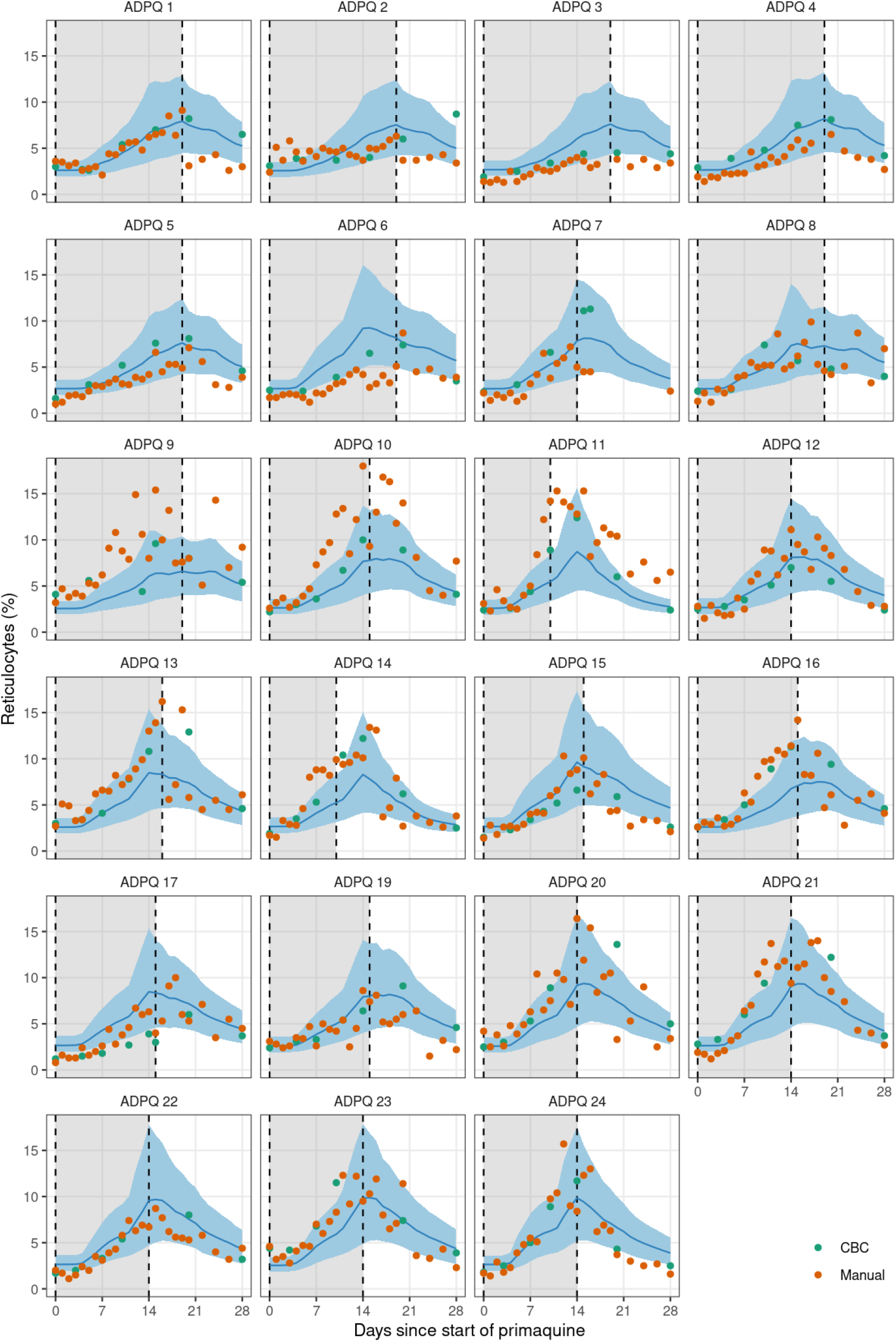
Leave-one-out predictions for the reticulocyte percentages over time for each individual in the ascending-dose study. Dots show the reticulocyte measurements (green: CBC; orange: manual reading). Blue lines (shaded areas) show the mean model prediction (90% predictive intervals). The grey shaded area indicates the primaquine dosing period.

The model was then used to predict the outcome of giving a single 45mg primaquine dose for the 16 individuals who participated in the second study (Figure 1d-e). The model predicted out-of-sample that the nadir haemoglobin would occur on day 4, whereas the observed nadir was day 6. The model predicted a substantial decrease in haemoglobin on day 4 (2.4g/dL), when there was a near-maximal reduction in RBC lifespan despite the effective dose being only 50% of the administered 45mg dose. This daily decrease was larger than those observed (Figure 1d), and as a consequence, the model over-predicted the increase in reticulocyte production (as this is primarily driven by the rate of haemolysis). This resulted in an over-estimation of the reticuloctye percentage on day 13 (Figure 1e). When fitting the model to both datasets jointly (ascending dose regimens and single dose), the model could not capture the exact haemoglobin and reticulocyte kinetics in both scenarios, indicating a problem with the parameterisation of the hysteresis in effect.

The model estimated that in these non-anaemic healthy G6PD deficient adult males, the major driver of increased erythropoeisis was the rate of haemoglobin decline rather than the absolute fall (absolute decrease from baseline). Peak reticulocytosis occurred approximately at the same time as the observed haemoglobin nadir in most volunteers. This could only be fitted to the model by adding as an explanatory covariate in the bone marrow production function: the change in haemoglobin over the previous 24 hours.

13 volunteers (50%) participated in both studies with over one year in between. In theory, these healthy participants should have had the same fundamental parameters (e.g. steady state haemoglobin, expected red cell lifespan, and primaquine dose response). However, several participants had considerably different baseline haemoglobin values and markedly different responses to primaquine were observed. For example, subject 11 who was a notable outlier in terms of primaquine-induced haemolysis in the ascending dose study, had a fall of only 3.4g/dL following the single dose (almost 3 years later), less than expected under the model. His baseline haemoglobin was nearly identical in Parts 1 and 2 (14.7g/dL and 14.5g/gL, respectively) but the reticulocyte count was lower in the single dose study (2.4% and 1.4%, respectively).

### Primaquine-induced red cell age reduction

The primary aim of the within-host model was to estimate the relationship between primaquine dose (in mg/kg base equivalent) and the reduction in the circulating red cell lifespan. If this can be estimated reliably then generalisable predictions can be made regarding the effect of a specific dose regimen. The mean steady state erythrocyte lifespan in these G6PD deficient volunteers was estimated at approximately 60 days (95% CrI: 55 to 65), with 80% of individuals having an estimated red cell lifespan between 38 and 82 days. These estimates are considerably shorter than for G6PD normal individuals (usually quoted as approximately ∼120 days [14]), as evidenced by the high mean reticulocyte counts at enrolment. This indicates a higher average erythrocyte turnover in G6PD deficiency [11].

Figure 5 panels A and B show the estimated delay in effect and dose-response under the model fitted to the ascending-dose regimen data. If a fixed daily dose was given, it would take approximately one week to reach maximal effect. Near maximal effects are achieved with doses of 0.4mg/kg or above (25mg in a 60kg adult), although there is considerable inter-individual variation both in the slope and the maximal effects estimated across the participants. Subject 11, who was a clear outlier in his haemolytic response to primaquine, had an estimated 60% reduction in circulating erythrocyte lifespan, from 52 days (95% CrI: 47 to 56) to 31 days (95% CrI: 27 to 36).

**Figure 5:**
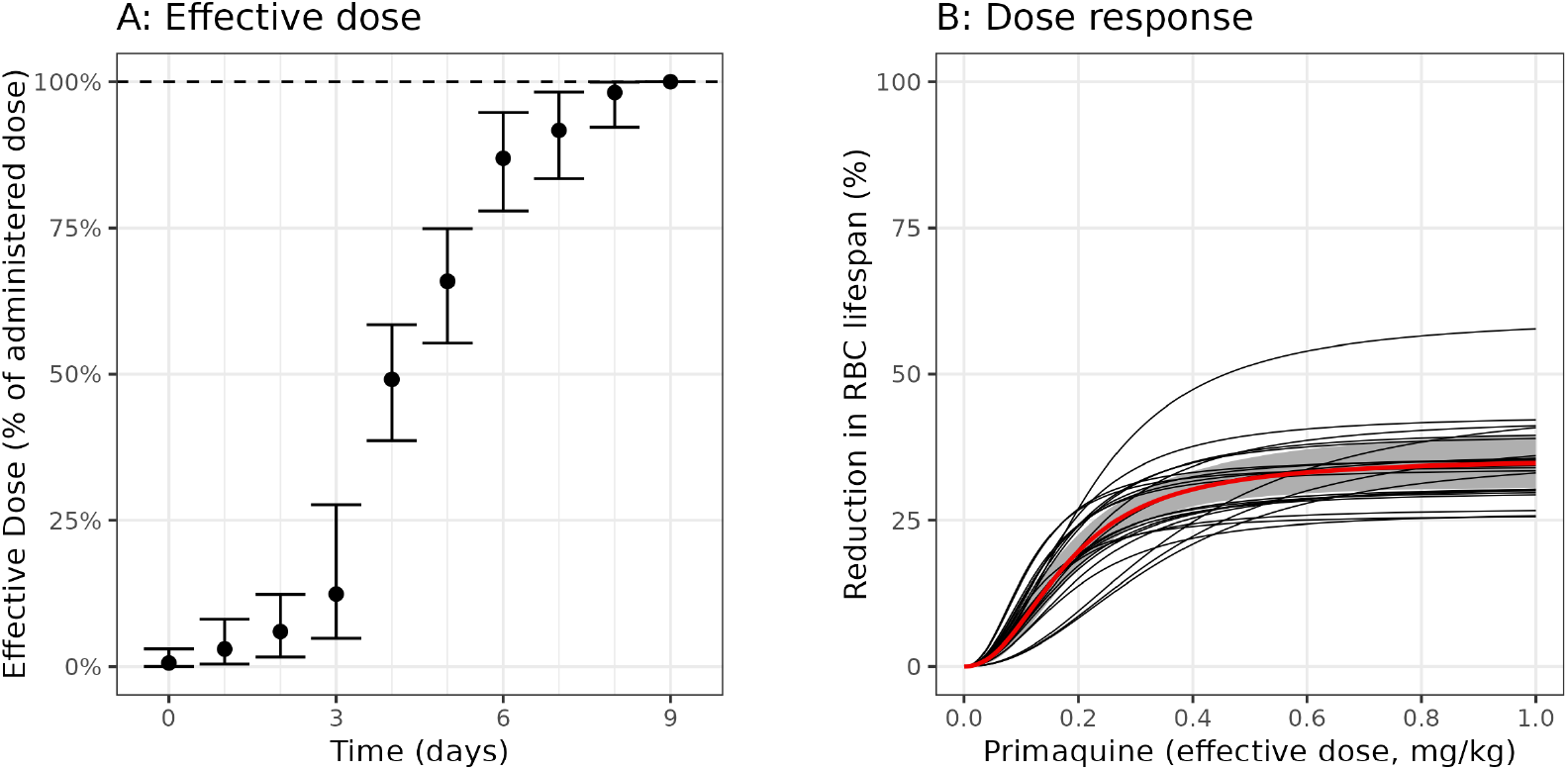
Posterior estimate of the pharmacodynamic model of primaquine-induced haemolysis. Panel A shows the delay in reaching the full effect when administering a fixed daily primaquine dose. Bars show 95% credible intervals (CrI). Panel B shows the relationship between the effective dose and the reduction in the erythrocyte lifespan. The black lines show mean posterior dose-response curves for each of the 23 participants; the red line (grey shaded area) shows the population mean response (95% CrI).

### Optimal ascending dose regimen

Total primaquine dose is the primary determinant of radical curative efficacy in relapsing malaria. A large individual patient data meta-analysis estimated that total doses between 5 and 7mg/kg achieved near maximal reductions in the relative risk of *P. vivax* recurrence over 6 months, irrespective of geographic origin and transmission intensity [15]. Based on this result, we used the within-host model to estimate how a target total dose of 5mg/kg could be administered optimally to a G6PD deficient hemizygote male over a period of 14 days or less. We defined optimal as the regimen which minimises the probability under the model that the individual will have a haemoglobin drop of *>*1g/dL in a single day. Figure 6 shows the estimated optimal ascending dose regimens for 5mg/kg administered over 10 or 14 days. It is not possible to administer 5mg/kg total dose over 10 days safely. Nearly 50% of individuals are predicted to experience a maximal daily fall *>*1g/dL. However, 5mg/kg can be administered safely over 14 days, allowing time for additional compensatory erythropoesis. The estimated optimal ascending dose regimen is similar to regimens given to some participants in the healthy volunteer trial, albeit with slower increases at the start and faster increases at the end of the two week regimen. Under this regimen, only 3% of individuals are predicted to have maximal daily falls over 1g/dL. For an individual with a baseline haemoglobin of 15g/dL, the mean predicted overall drop in haemoglobin is 4.4g/dL (95% predictive interval: 2.7 to 6.5g/dL), with the nadir haemoglobin occurring between days 13 and 17.

**Figure 6:**
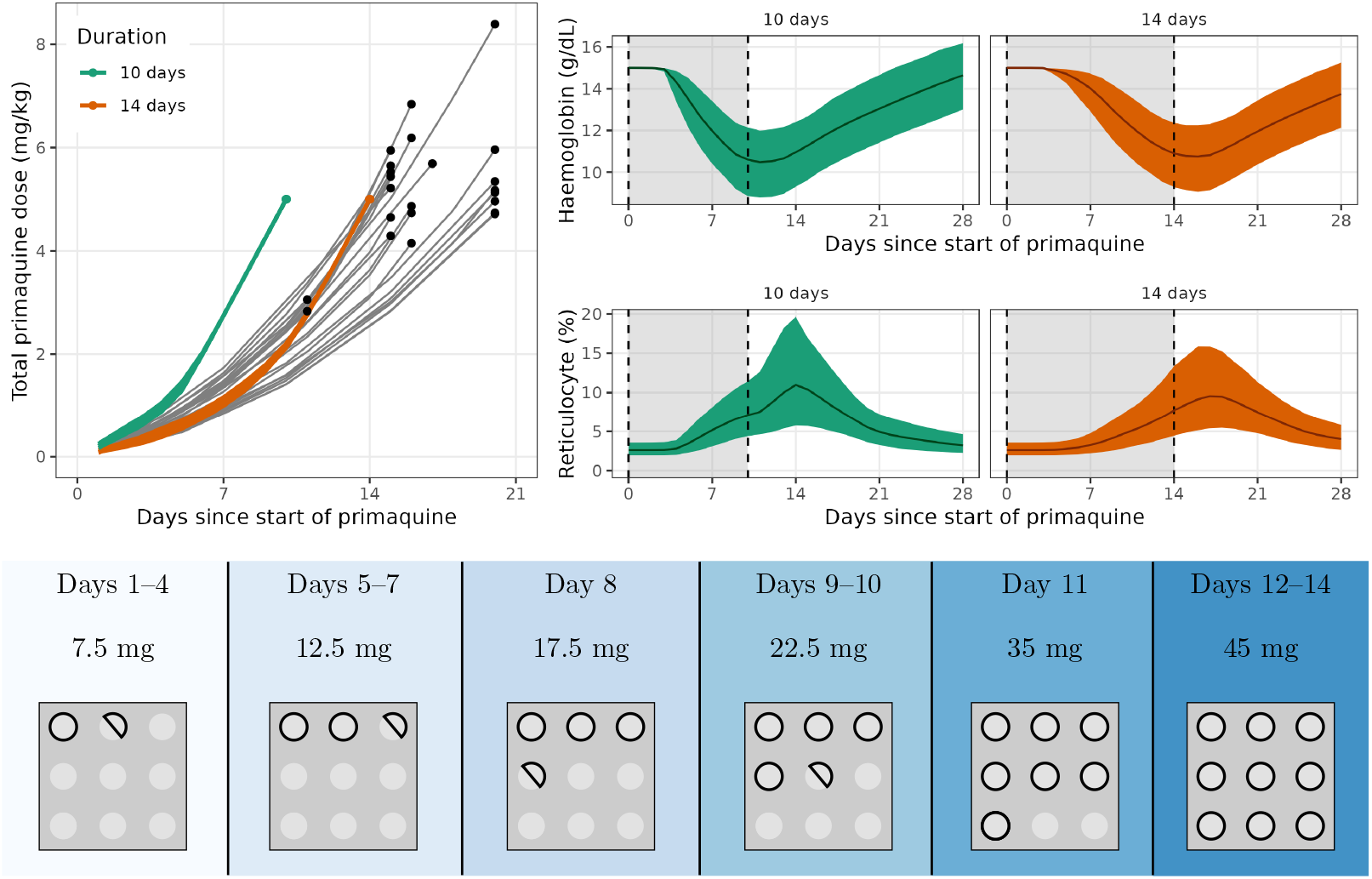
The optimal 10-day and 14-day primaquine ascending dose regimens. Top left: A comparison of the optimal regimens (coloured lines) to the regimens administered to each participant in the ascending dose study (grey lines). Points indicate the end of each regimen. Top right: The predicted daily haemoglobin for the optimal regimens (upper panel) and the predicted daily reticulocyte levels for the optimal regimens (lower panel). Bottom: the predicted safest ascending dose regimen that comprises no more than 6 different dose sizes. This assumes tablet sizes of 5mg which can be halved. This regimen is almost as safe as the optimal regimen; only 4% of individuals are predicted to have maximal daily falls over 1g/dL (cf. 3%).

## Discussion

The highly detailed serial haemoglobin and reticulocyte data from the ascending primaquine dose study in healthy volunteers allowed for the development of a within-host model of red cell turnover and primaquine-induced haemolysis in G6PD deficiency. The model structure is based on a good understanding of the main underlying cellular mechanisms, elucidated from clinical studies conducted over 50 years ago [7, 16]. An important result is the characterisation of the substantial intra- and inter-subject variability in haemolytic response. It is well known that G6PD levels vary substantially both within and between individuals. The high basal reticulocyte counts indicate that under normal conditions G6PD deficient subjects have increased erythrocyte turnover. Presumably, varying environmental and nutritional oxidant stresses result in variation in the proportion of the erythrocyte population which is susceptible to further (iatrogenic) oxidant haemolysis. Patients who are anaemic as a result of malaria have already lost their most oxidant sensitive (i.e. older) erythrocytes [17]. Thus the reductions in haemoglobin that result from primaquine oxidant haemolysis in G6PD deficient patients with acute malaria are expected to be less than those observed in this study of healthy subjects. This is important as the main life-threatening dangers of acute haemolysis are haemoglobinuric renal failure (proportional to the fall in haemoglobin) and severe anaemia (depends on the fall, the initial haemoglobin concentration, and the chronicity of any preceding anaemia). Despite substantial intra- and inter-individual variation, a mathematical model based upon the known haemolytic pharmacodynamics fitted well to the documented changes in haemoglobin and reticulocyte counts in the ascending dose primaquine challenged G6PD deficient volunteers. We show that a total dose 5mg/kg of primaquine, which is sufficient for radical cure in most patients in endemic areas [15], can be administered safely over 2 weeks to subjects with moderate severity G6PD deficiency. Only a minority of individuals are predicted to have large daily falls in haemoglobin under this regimen. If a 2 week radical curative regimen was shown safe in G6PD deficient patients with vivax malaria, this would be a substantial improvement on the current 8-weekly regimen, affecting the treatment of over 10% of individuals in areas where G6PD deficiency alleles are common [18].

Although potentially dangerous in G6PD deficient individuals, billions of individuals have been given primaquine over the past seventy years, with no G6PD deficiency testing in the majority [19, 20]. Primaquine has been used in mass treatments in areas with a high prevalence of *G6PD* deficiency mutations [21]. Primaquine-induced haemolysis is inevitable in G6PD deficiency, but it can be controlled. If iatrogenic haemolysis is balanced by erythropoiesis, the red cell population becomes progressively younger (and more resistant to oxidant stresses) with only a modest and temporary anaemia. This is because haemolysis is greater in older erythrocytes. G6PD deficiency reflects an unstable enzyme which degrades more rapidly than in normal subjects. The different inherited genetic *G6PD* variants encode enzymes which vary in their stabilities or catalytic activities, affecting the proportion of circulating erythrocytes which are vulnerable to oxidant haemolysis [22]. The resulting haemolysis ranges from less severe in African A-variants to more severe in the common Mediterranean variant. However, within each genotype there is substantial phenotypic variability so serious haemolysis can occur even with the less severe variants [22]. Titrating primaquine dosing against haemolysis reduces the risk of dangerous haemolytic anaemia.

The choice of a radical cure dosing regimen in G6PD deficiency depends on many factors. First, the benefits of preventing relapse must outweigh the risk of haemolysis [10]. The prevalence of the different *G6PD* genetic variants and their associated degree of G6PD deficiency varies substantially [1], and vivax relapse rates also vary from approximately 20% to nearly 100% [15, 23]. Second, there is wide intra- and inter-individual variability in haemolytic responses, requiring a reasonably wide margin of safety. Third, complex drug regimens may result in poor adherence, and could risk unintentional overdosing. Fourth, the longer the duration of treatment, the less likely it will be completed [24].

A key limitation of the primaquine-induced haemolysis red blood cell model is the lack of adequate characterisation of hysteresis in the pharmacodynamic effect. Primaquine is a pro-drug for which both the parent compound and the active metabolites are rapidly eliminated (half-life *<*12 hours). However, there is a long delay between drug administration and haemolysis, as shown in the single dose study (nadir haemoglobin occurs almost one week later) [11]. Our model is phenomenological with respect to the delay in effect. We do not mechanistically model this delay in effect, which is driven by depletion of the erythrocyte’s G6PD mediated antioxidant defence. The non-parametric weighting of previous doses providing an ‘effective dose’ captures the profiles observed in the ascending dose data but does not fully capture the single dose data. In addition, there is considerable intra- and inter-individual variability in haemolytic response. We do not have a good biomarker predictive of this variability. Intra- and inter-individual variability may be driven by differences in recent haemolytic insults, for example from diet or infection. Our model assumes that at enrolment the distribution of circulating red cell ages is approximately uniform. Deviations from this assumption have important consequences on the impact of primaquine administration. Most importantly, this limits the ability to predict primaquine-induced haemolysis reliably in patients with vivax malaria. Malaria causes haemolysis of non-parasitised older red cells (which are removed by the spleen), this would be expected to attenuate the effect of primaquine [17]. However, *P. vivax* has a tropism for younger reticulocytes, and in some patients with haematinic deficiencies there may be an attenuated bone marrow response. Extrapolating from healthy volunteers to malaria patients is uncertain and data from patients are now needed to calibrate the model.

This model allows prediction of the haemolytic dose-response relationship in subjects with Southeast Asian variants of G6PD deficiency. This should be applicable to other variants of similar severity and will overestimate haemolysis in milder variants. It cannot be extrapolated directly to more severe variants (e.g. *G6PD* Mediterranean). However, the large variation of phenotypes within genotypes means that even individuals with less severe variants may occasionally experience severe haemolysis. More information is needed to calibrate these risks. The proposed ascending dose regimen should now be tested for safety in patients with *P. vivax* malaria.

## Material and Methods

### Clinical study

The Primaquine Challenge study was a two-part regimen-adaptive open-label study that aimed to assess the safety and tolerability of ascending primaquine dose regimens in healthy Thai male G6PD deficient volunteers. Clinical details have been published previously [11]. All volunteers provided fully informed written consent and agreed to all study procedures. The two parts of this study were approved as separate studies. Both parts were approved by the Faculty of Tropical Medicine’s Ethics Committee (MUTM 2017-036-01 and MUTM 2021-031-02) and the Oxford Tropical Research Ethics Committee (OxTREC, number 48-16). The study protocols were pre-registered on the Thai Clinical Trial Registry (TCTR, numbers TCTR20170830002 and TCTR20220317004).

In brief, the study subjects were healthy male adult volunteers (18 to 65 years of age) recruited in Bangkok (Faculty of Tropical Medicine, Mahidol University) with G6PD deficiency confirmed by a validated quantitative spectrophotometric G6PD assay (enzyme activity *<* 30%), and with a known genotype. Individuals with the G6PD Mediterranean variant (C563T) or any previously uncharacterised mutations were excluded from the study. A total of 27 individuals were recruited over four years (the study was interrupted substantially by the COVID-19 pandemic). The study had two parts. In Part 1, ascending primaquine doses over 15 to 20 days were given (daily doses ranging between 7.5 and 45mg base). In Part 2, single 45mg doses were given. 24 individuals participated in Part 1. COVID-19 mitigation measures interrupted the end of Part 1 and considerably delayed finishing the study. 13 subjects from Part 1 also participated in Part 2, and 3 additional subjects were enrolled to capture inter-individual variability better following the single 45mg dose.

All subjects were admitted to the phase 1 unit at the Hospital of Tropical Diseases during the period of drug administration (between 15 to 20 days) for Part 1, and for one week following the single dose for Part 2. All primaquine doses were directly observed. In Part 1, follow-up occurred over 49 days following the first primaquine dose; and in Part 2 over 14 days. Venous blood samples for complete blood counts (CBC) were taken at screening and then every 4-5 days during drug administration. Fingerprick blood samples were taken daily during drug administration and at follow-up visits for manual reticulocyte count readings and Haemocue haemoglobin measurement. The primaquine regimens administered were adapted throughout the trial based on accrued data, perceived safety by the study investigators and using a set of pre-specified rules [11]. The aim of Part 1 was to determine a regimen that resulted in a steady decrease in haemoglobin during primaquine dosing with a total decrease which was less than 40% of baseline; this haemoglobin drop was then compared with that resulting from a single high dose (45mg base equivalent) primaquine (Part 2).

### Compartmental model of red blood cell dynamics

Red blood cells are produced through a highly regulated process (erythropoiesis), which starts with the commitment of haemopoietic precursors and the generation of immature nucleated red cells (normoblasts) in the bone marrow. These normoblasts lose their nuclei and mature into reticulocytes, which are then released into the bloodstream. These cells lose their ‘reticulated’ appearance (resulting from residual mRNA) after 1-2 days and become mature erythrocytes. At steady state in normal adults, reticulocytes constitute approximately 1% of all circulating red blood cells. Loss of red blood cells (in this case from haemolysis) triggers a feedback response resulting in increased production in the bone marrow and an earlier release of reticulocytes into circulation [16]. An increased reticulocyte count can thus reflect either haemolysis that has just occurred (i.e. reticulocytes are a greater proportion of all circulating red blood cells), a recent increased bone marrow production, or both [16]. Reticulocyte counts are thus an important proxy marker of bone marrow erythropoietic activity.

In this work we update a previously published model of red blood cell dynamics [12] to infer the dose-dependent haemolytic effect of primaquine in G6PD deficiency, and characterise the dynamics of red blood cell production and response to haemolysis. The key underlying model assumption is that primaquine-induced haemolysis is red cell age-dependent: i.e. the older cells haemolyse first. This is parameterised in the model as a dose-dependent reduction in the red blood cell lifespan.

#### Steady state dynamics

We denote values at steady-state with the superscript *, e.g. the steady state haemoglobin concentration is denoted Hb*. The model captures variation over time in haemoglobin concentrations and in the proportion of circulating red cells which are reticulocytes, indexed by *t*. We assume that at the first time point *t* = 0 (i.e. before drug administration) the subject is at steady state, Hb_*t*=0_ = Hb*. Steady state means that over one day the number of red cells which are removed from the circulation equals the number liberated (and thus newly produced) by the bone marrow. The model is parameterised in terms of the number of circulating red blood cells (denoted C_*t*_), assuming a constant one-to-one relationship between the number of circulating red blood cells and the observed haemoglobin concentration. This assumes no changes in mean corpuscular haemoglobin content. At *t* = 0, Hb_*t*_ = ΛC_*t*_. We note that the dynamics of the system are invariant to the value of the scaling factor Λ so we use a computationally convenient value.

The numbers of red blood cells at the different stages are modelled using non-linear difference equations broken into daily blocks. The number of red blood cells in the *i*^th^ block of each of the three stages of red blood cell maturation at time *t* is denoted by 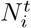, *i* = 1, …, *T*_*N*_ for normoblasts; 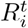, *i* = 1, …, *T*_*R*_ for reticulocytes; and 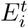, *i* = 1, …, *T*_*E*_ for erythrocytes. *T*_*N*_, *T*_*R*_, and *T*_*E*_ are the maximal survival ages in units of days for normoblasts, reticulocytes, and erythrocytes, respectively. For example, 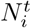 corresponds to the number of normoblasts that are *i* days old in the system at time *t*. Normoblasts are contained within the bone marrow (we assume no liberation). Reticulocytes are formed in the bone marrow and then emerge into circulation at a varying age, Transit(Hb_*t*−1_), which we model as a function of the haemoglobin concentration at the previous time-point [16]. At steady state in a normal individual, reticulocytes emerge from the bone marrow after approximately three and a half days (i.e. Transit[Hb*] = 3.5 days) and spend approximately one and a half days in circulation before becoming mature red blood cells (erythrocytes). An overview of the model structure is shown in Figure 2.

#### Release time of reticulocytes

At time *t*, the total number of circulating cells is given by:

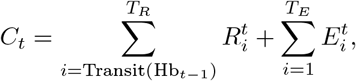

The quantity 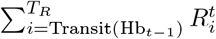 is the total number of circulating reticulocytes. The proportion of reticulocytes in the circulation is then simply 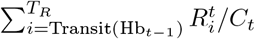. Note that this assumes Transit(·) takes integer values only; in the more general case we take the fractional part of the first term in the sum.

The transit time of reticulocytes into circulation is parameterised as a simple exponential function:

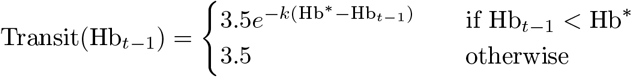

where *k* is the slope coefficient which determines how quickly the transit time reduces as the haemoglobin falls below steady state; 3.5 is the steady state value (days).

#### Production of normoblasts

Haemolysis triggers an increase in production of red blood cells in the bone marrow. This is captured by the ‘production function’ *ρ* (expressed as a fold change relative to steady state). We parameterise *ρ* as a function of both the absolute difference in haemoglobin from steady state (i.e. total loss of red blood cells relative to steady state) and the change in haemoglobin compared to the previous day (reflecting the rate of haemolysis). For both components, we use second degree polynomials:

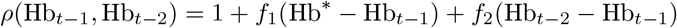

where for *s* = 1, 2:

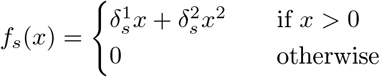

#### Forward time simulation

The model iterates daily the production and release time processes to determine changes over time in haemoglobin concentration and reticulocyte counts for a given set of parameters. At time *t* = 0 the model is initialised at steady state:

- 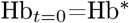
- 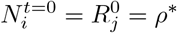 for all *i* = 1, …*T*_*N*_, *j* = 1, …, *T*_*R*_
- 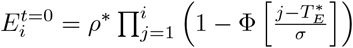, where *i* = 1, …*T*_*E*_

In each subsequent time step 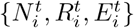 are updated from 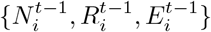 using the following difference equations:

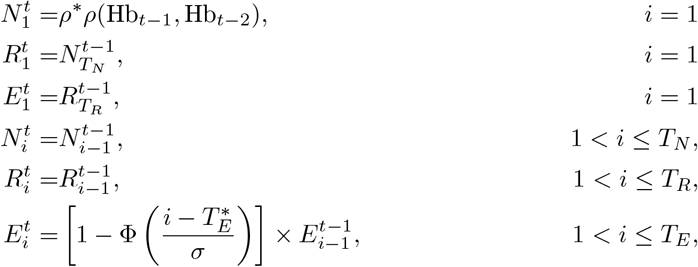

The value *ρ*\* is an arbitrary scalar which determines the relationship between the total number of circulating red blood cells and the units of haemoglobin (i.e. determines the value of Λ); Φ is the cumulative standard normal distribution (probit function); 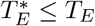 is the expected red blood cell lifespan for the individuals. The function 1 − Φ parameterises the steady state age-dependent removal of older red blood cells as a sigmoid process. The parameter *σ* determines the ‘steepness’ of this removal process when the age approaches 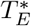. The probit curve results in negligible decay until 3-4 standard deviations (*σ*) from the mean value, which in absence of drug is 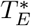. Thus at steady the number of erythrocytes is approximately uniformly distributed over the interval 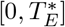.

#### Primaquine-induced haemolysis in G6PD deficiency

Primaquine administration results in dose-dependent haemolysis in G6PD deficient individuals. This represents loss of older red blood cells which contain lower G6PD enzymatic activity, and are therefore susceptible to primaquine-induced oxidative stress [7]. Delay between starting primaquine and the onset of haemolysis is attributed to depletion of the remaining intraerthrocytic anti-oxidant defences in the oldest erythrocytes. Primaquine is not the active moiety - it is converted to bioactive intermediates which cause oxidative haemolysis and the beneficial antimalarial effect. This is relatively rapid (although the degree of conversion varies between individuals), so for the purposes of this model, the primaquine dose is considered a surrogate for the amount of oxidative intermediates produced. Our model assumes oral daily primaquine administration. When modelling the time interval [0, *T*_max_], we represent the primaquine regimen as a vector 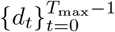, where *d*_*t*_ is the mg/kg dose administered on day *t*. Primaquine is metabolised rapidly and so can be modelled as daily ‘pulses’. However, a complicating factor is the delay between the primaquine dose administration and the resulting haemolysis (hysteresis).

##### Effective dose: delay in effect

We model hysteresis in the haemolytic effect of primaquine by defining an ‘effective dose’ *x*_*t*_ at time *t* which is a function of the previously administered doses.

The effective dose *x*_*t*_ is a time-weighted average of the previously administered doses:

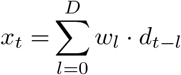

We used a symmetric Dirichlet prior distribution for the weights **w** = (*w*_0_, *w*_1_, …, *w*_*D*_), with concentration parameter 1.

##### Dose-dependent reduction of red blood cell lifespan

We model the dose-dependent haemolytic effect of primaquine by defining the reduced erythrocyte lifespan at time *t* to be *T*_*E*_(*x*_*t*−1_), a function of the ‘effective dose’ *x*_*t*−1_ from the previous day. Because the effective dose is a weighted average of previous doses, this takes into account the delay in the effect. The dose-dependent effect of primaquine is modelled as an E_max_ function of the effective dose *x*:

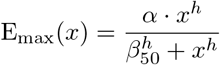

where *α* ∈ [0, 1] is the maximum achievable primaquine-induced decrease in relative lifespan. The parameter *β*_50_ is the half-maximal effect dose, i.e. E_max_(*β*_50_) = *α/*2. The parameter *h* is the ‘Hill’ coefficient which determines the slope of the dose-response curve.

The reduced red blood cell lifespan at time *t* is thus:

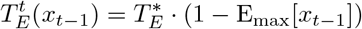

Therefore, at each time step the number of erythrocytes is updated with the following difference equation:

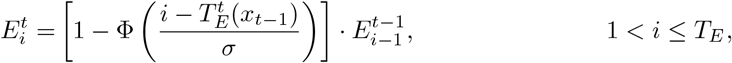

### Optimal ascending dose regimen

An ideal primaquine regimen for individuals with G6PD deficiency would provide a radical curative dose, over a reasonable period (2 weeks as for the standard regimen), whereby falls in haemoglobin are gradual [12]. A review of the available evidence suggested that a total dose of 5mg base/kg provides near maximal anti-relapse (radical cure) activity [15]. We defined ‘optimal’ as the regimen which minimises the probability of a fall *>*1g/dL over the course of a single day.

We used the posterior distribution over the model parameters when fitted to the ascending dose study data to identify optimal 10-day and 14-day regimens, subject to the following criteria:

- Each daily dose was a multiple of 2.5mg (equivalent to half of a 5mg tablet);
- Each daily dose was at least as large as the previous dose (i.e., ascending); and
- The total dose was 300mg (equivalent to 5mg/kg for a 60kg individual).

There were 78,796 possible 10-day regimens, and 3,648,057 possible 14-day regimens that satisfy these criteria. We used a brute-force approach, and evaluated each candidate regimen by simulating the model forward from time *t* = 0 for 1,000 virtual individuals. We drew 1,000 posterior samples for the population model parameters and participant-specific model parameters (individual random effects). Each individual was assigned a random body weight which was normally distributed with mean 60 and standard deviation 5. For each individual, we recorded the maximum daily drop in haemoglobin, and scored each candidate regimen by the proportion of individuals who experienced a drop of *>* 1g/dL.

### Model fitting

Model parameters were estimated under a Bayesian hierarchical model framework allowing for inter-individual variation in the haemolytic effect of the drug, the steady state red blood cell lifespan, the steady state haemoglobin, bone marrow response to haemolysis, and reticulocyte response to haemolysis (transit time from bone marrow to circulation).

#### Data notation

In all the following the subscript *t* denotes the time in days, and *i* denotes the subject number. The model is fitted to four types of data: (i) CBC haemoglobin values, denoted 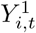; (ii) Haemocue® haemoglobin values, denoted 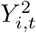; (iii) CBC reticulocyte values, denoted 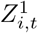; and (iv) manually read reticulocyte values, denoted 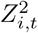.

The residual errors for the modelled haemoglobin concentrations were assumed to be normal; for the reticulocyte counts, they were assumed to be log-normal.

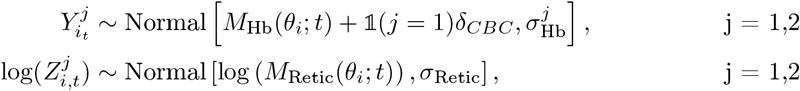

The parameter *δ*_CBC_ adjusts for the systematic differences in the haemoglobin measurement when between CBC and Haemocue [11]. *M*_Hb_(*θ*_*i*_; *t*) and *M*_Retic_(*θ*_*i*_; *t*) correspond to the model predicted haemoglobin concentration and reticulocyte counts for a given individual-specific parameter vector *θ*_*i*_ = *θ* + *η*_*i*_; *θ* is the set of population-level parameters:

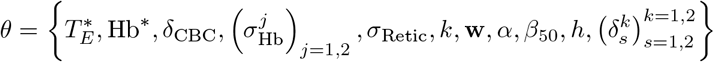

*η*_*i*_ are the individual random effects which capture variation between individuals. *η*_*i*_ has dimension 8. Individual random effects are added to all parameters except for 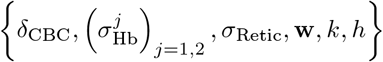. The prior distribution for the individual-specific parameters was assumed to be multivariate normal with mean zero and covariance matrix Ω. Ω was decomposed into a vector of standard deviation parameters and a correlation matrix with a Cholesky prior distribution with shape parameter equal to 2 [25, 26]. The prior distributions for all parameters, as well as all fixed model parameters, are provided in Table S3 along with their interpretation.

Analysis was conducted in R 4.1.3 [27] and model fitting was performed using Stan 2.34 [25] and the CmdStanR package 0.7.1 [28]. Four chains were implemented and 1,000 posterior samples were retained from each chain after a burn-in of 1,000 iterations, resulting in 4,000 samples per parameter for the calculation of posterior summaries. The posterior summaries calculated for each parameter were the mean of these samples and the 90% credible interval, which was calculated from the 5^th^ and 95^th^ percentiles of samples. 80% predictive intervals to quantify inter-individual variation in Table 2 were derived from the 10^th^ and 90^th^ percentiles of the draws for the individual-specific parameters.

Convergence was checked from trace plots and standard MCMC diagnostic criteria including the 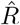 statistic (*<*1.05 was considered to demonstrate acceptable convergence), and the effective sample sizes (ESS). Prior and posterior distributions were compared visually for the main parameters (Supplementary Figure S2).

A total of 1,000 random samples were drawn from population red blood cell model parameters (*θ*) derived from the posteriors to compare different dosing regimens. Every one of the red blood cell vector of parameters corresponds to a single red blood cell profile data set, where each of these vectors and a dosing regimen of primaquine, were input into the red blood cell model in order to simulate 1000 haemoglobin concentration and reticulocyte counts. We draw 1000 random sets of participants specific red blood cell model parameters to simulate 1000 hypothetical red blood cell profiles (haemoglobin concentration and reticulocyte counts) and to compare alternative dosing regimens of primaquine to the current regimen administered by each individual. The process outlined above was repeated for each dosing regimen.

## Data Availability

All data and code are available on GitHub: https://github.com/jwatowatson/Primaquine-Challenge

## Code and data availability

All data and code are available on GitHub: https://github.com/jwatowatson/Primaquine-Challenge

## Acknowledgements

We are very grateful to the volunteers who participated in this study. We thank the staff of the Clinical Therapeutics unit and the laboratories in the Hospital for Tropical Diseases, Bangkok, which provided essential monitoring. We thank Penporn Penpitchaporn for doing all manual reticulocyte count readings.

This study was funded by the MRC “Assessing the tolerability of a potentially safer radical curative regimen of primaquine in healthy volunteers with glucose 6 phosphate dehydrogenase” (MR/R015252/1). This UK funded award is part of the EDCTP2 programme supported by the European Union. JAW is a Sir Henry Dale Fellow funded by the Wellcome Trust (223253/Z/21/Z). NJW is a Principal Research Fellow funded by the Wellcome Trust (093956/Z/10/C). JAS is funded by an Australian NHMRC Leadership Investigator Grant (1196068), and RJC a NHMRC Emerging Leadership Investigator Grant (1194702).

This research was partly funded by Wellcome. A CC-BY or equivalent licence is applied to the author accepted manuscript arising from this submission, in accordance with the grant’s open access conditions.

---

**Figure S1:**
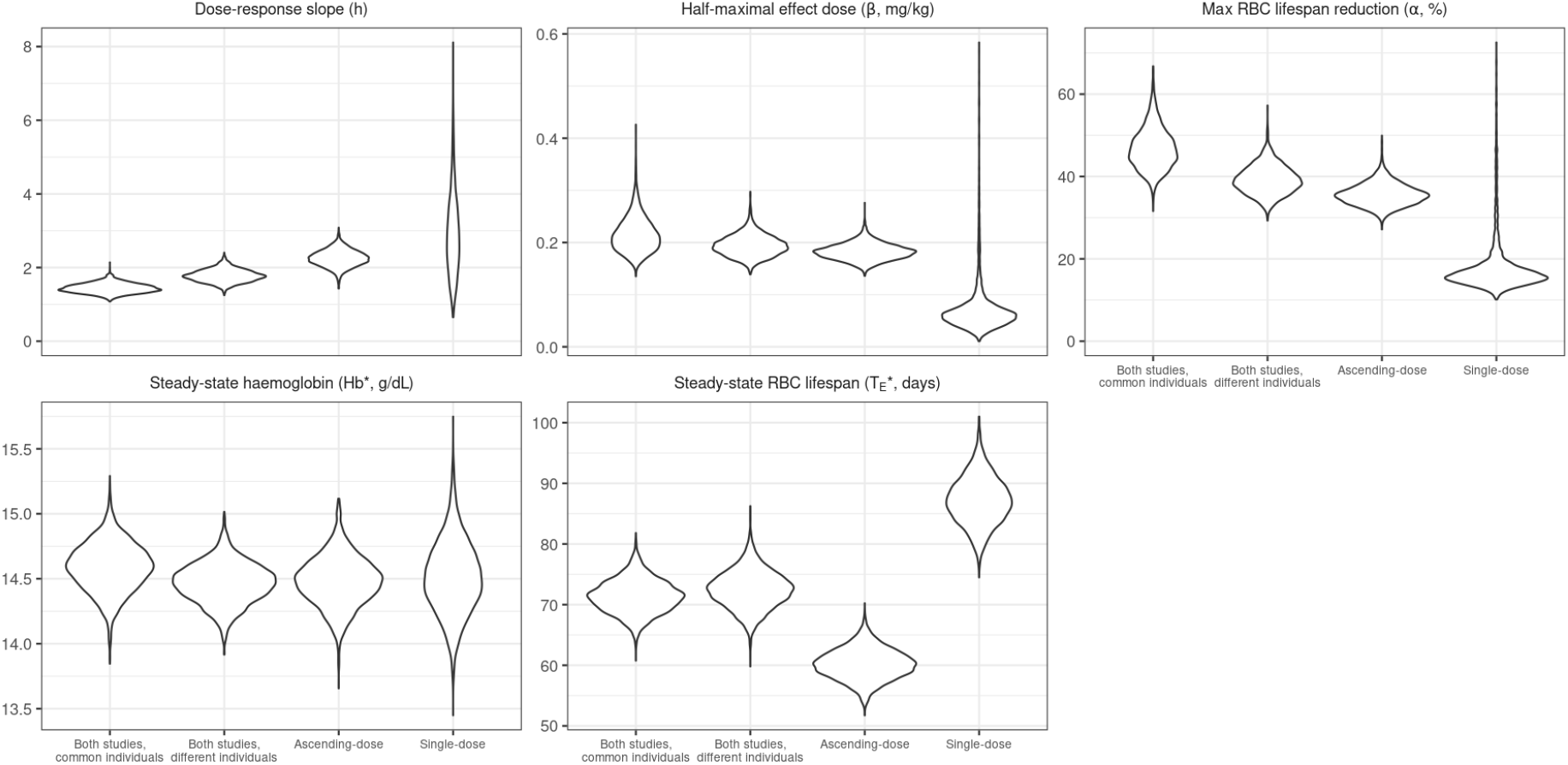
Comparison of the marginal posterior distributions for key model parameters when fitting to (a) both studies, assuming that individuals enrolled in both have identical parameters; (b) both studies, assuming that individuals enrolled in both have independent parameters; (c) only the ascending dose study; and (d) only the single dose study. There are clear differences between the dose responses and RBC lifespans estimated from the single dose study and from the ascending dose study. In particular, the single dose study estimates a much longer steady-state RBC lifespan, and a much smaller dose response that is triggered by very low doses.

**Figure S2:**
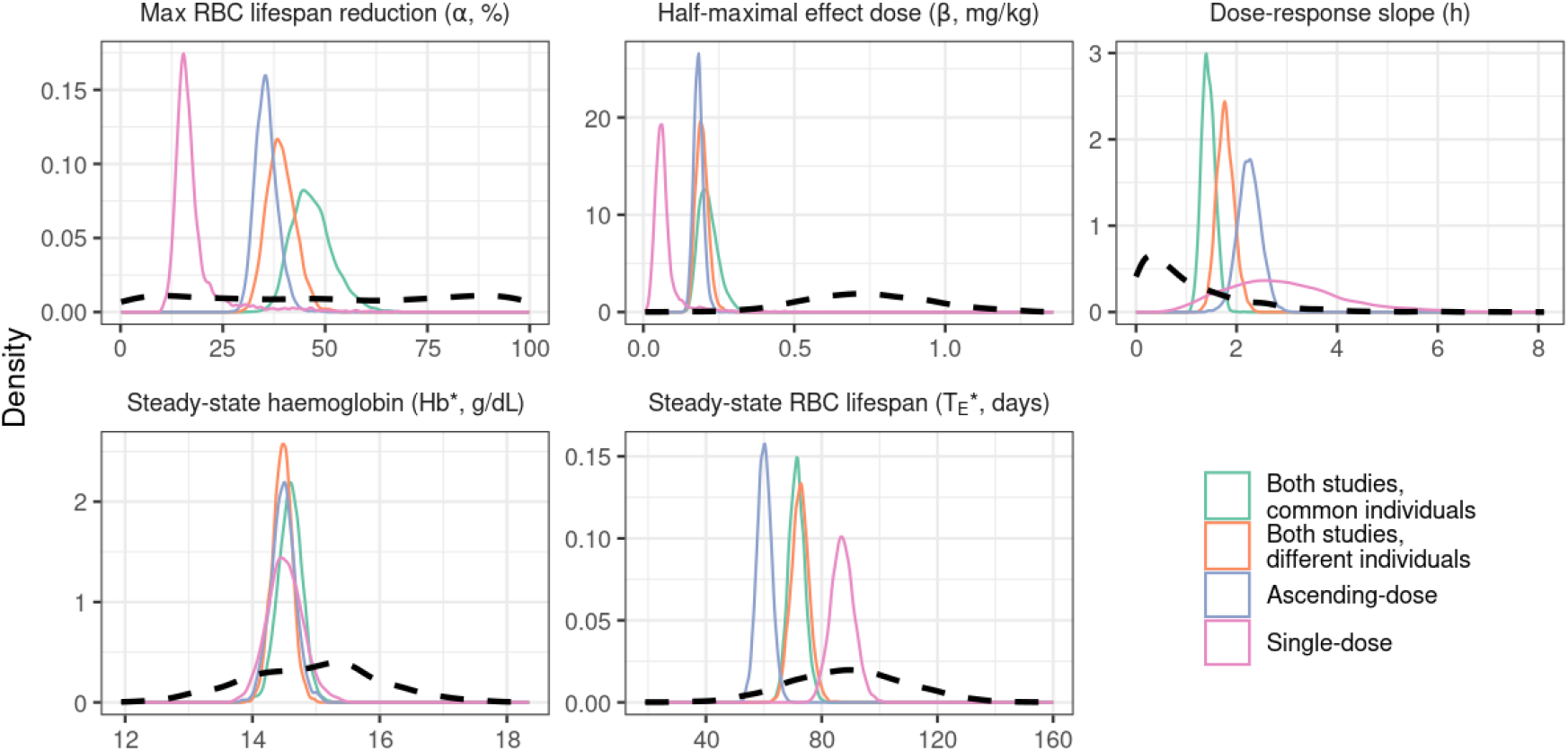
Comparison of the marginal posterior distributions for key model parameters to the prior distributions. Marginal posterior distributions are shown when fitting to (a) both studies, assuming that individuals enrolled in both have identical parameters; (b) both studies, assuming that individuals enrolled in both have independent parameters; (c) only the ascending dose study; and (d) only the single dose study.

**Figure S3:**
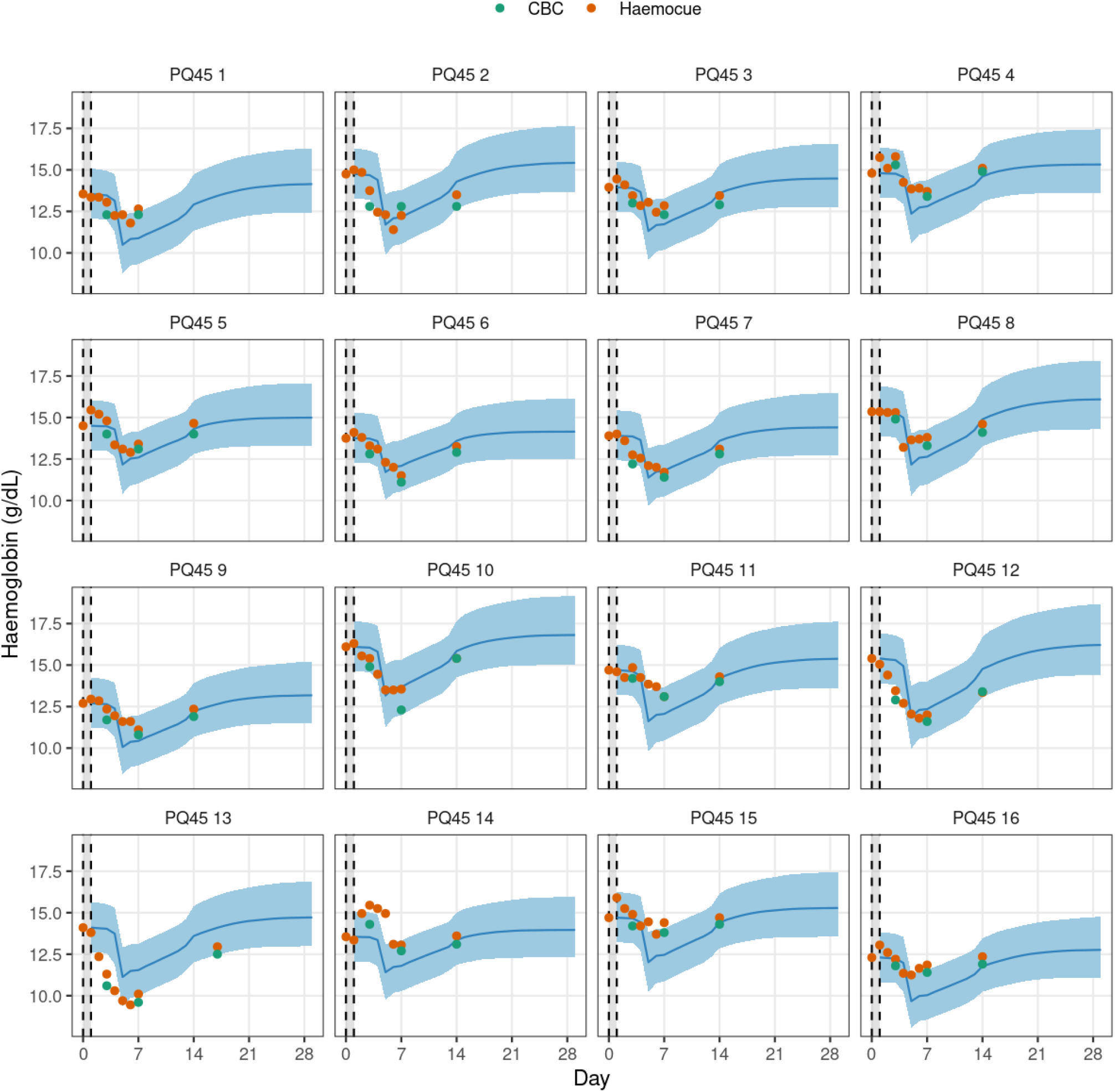
Haemoglobin predictions following the single 45mg dose, under the model fitted to ascending-dose study. The model predicts a rapid drop in Hb several days after the single dose; this is consistent with the data from some individuals (e.g., PQ45 2 and PQ45 10), but other individuals experienced a smaller, more gradual decrease in Hb (e.g., PQ45 1 and PQ45 11), an earlier drop in Hb (e.g., PQ45 12 and PQ45 13), or even no decrease in Hb (e.g., PQ45 14 and PQ45 15).

**Figure S4:**
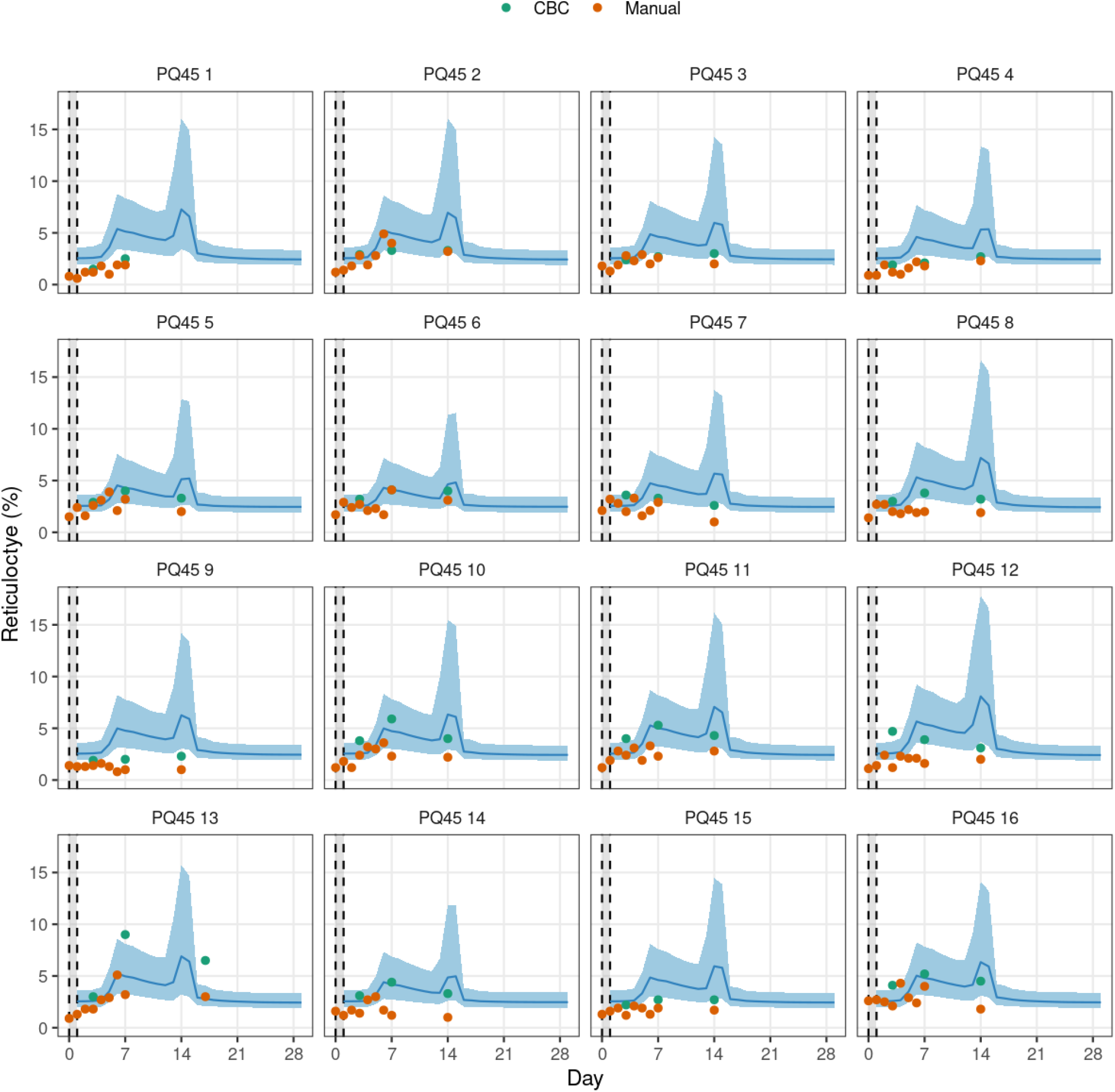
Reticulocyte predictions following the single 45mg dose, under the model fitted to ascending-dose study. The predictions are in reasonable agreement with the measurements taken over days 0 to 7, but the measurements taken on day 14 (day 17 for individual PQ45 13) do not provide evidence of a short, rapid increase in reticulocytes.

**Table S3:**
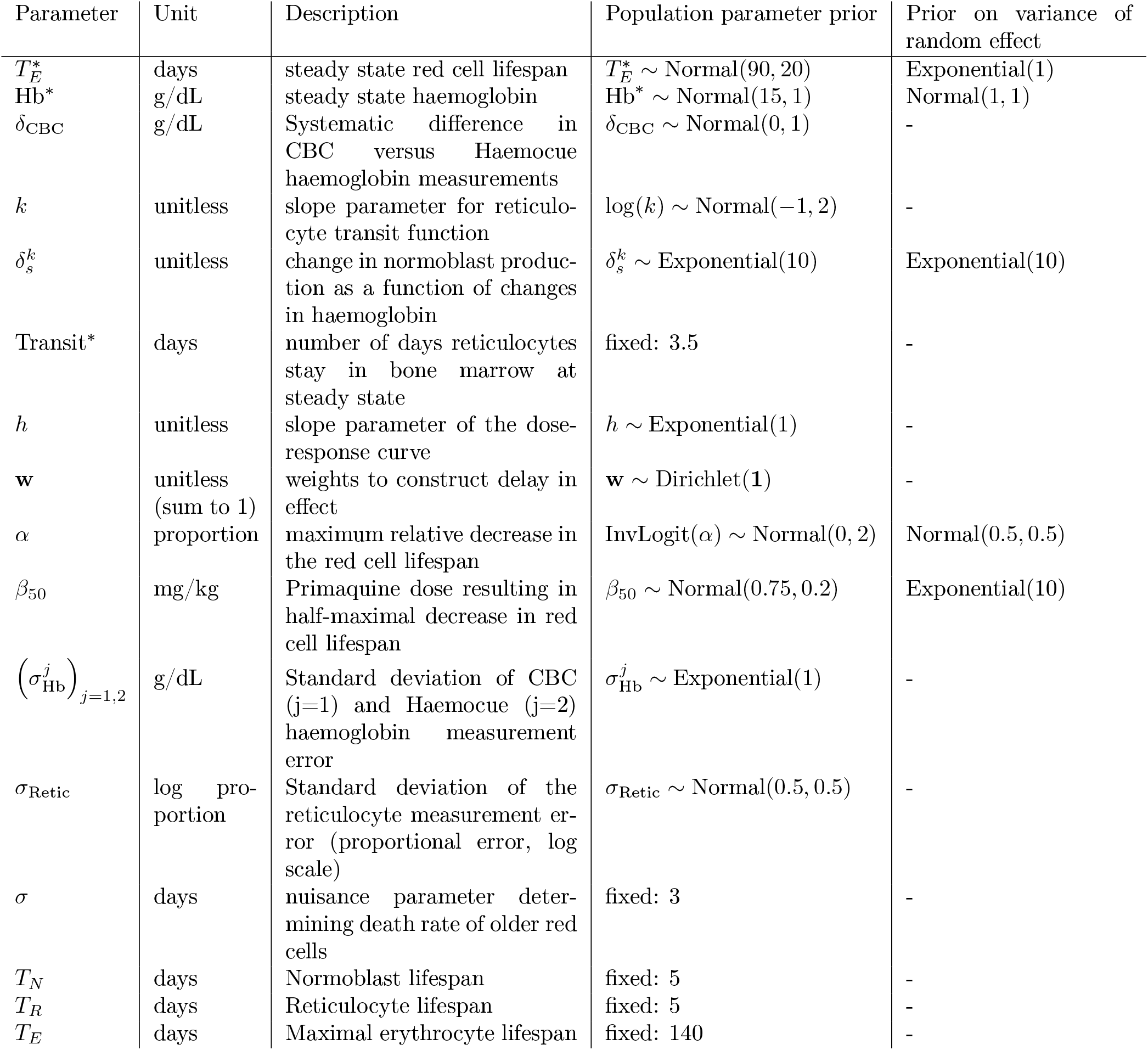
Prior distributions for model.

## References

[1] Rosalind E Howes, Frédéric B Piel, Anand P Patil, Oscar A Nyangiri, Peter W Gething, Mewahyu Dewi, Mariana M Hogg, Katherine E Battle, Carmencita D Padilla, J Kevin Baird, et al. G6PD deficiency prevalence and estimates of affected populations in malaria endemic countries: a geostatistical model-based map. PLoS Med, 9(11):e1001339, 2012.

[2] Ric N Price, Robert J Commons, Katherine E Battle, Kamala Thriemer, and Kamini Mendis. Plasmodium vivax in the Era of the Shrinking P. falciparum Map. Trends in Parasitology, 36(6):560–570, June 2020.

[3] J Kevin Baird. 8-aminoquinoline therapy for latent malaria. Clinical Microbiology Reviews, 32(4):10–1128, 2019.

[4] Saber Dini, Nicholas M Douglas, Jeanne Rini Poespoprodjo, Enny Kenangalem, Paulus Sugiarto, Ian D Plumb, Ric N Price, and Julie A Simpson. The risk of morbidity and mortality following recurrent malaria in Papua, Indonesia: a retrospective cohort study. BMC Med., 18 (1):28, February 2020.

[5] World Health Organization. WHO Guidelines for malaria. World Health Organization, 1st edition, 2023. URL https://www.who.int/publications/i/item/guidelines-for-malaria.WHO/UCN/GMP/2023.01Rev.1.

[6] Maria Domenica Cappellini and Gemino Fiorelli. Glucose-6-phosphate dehydrogenase deficiency. The Lancet, 371(9606):64–74, 2008.

[7] Ernest Beutler, Raymond J. Dern, and Alf S. Alving. The hemolytic effect of primaquine IV. The relationship of cell age to hemolysis. The Journal of Laboratory and Clinical Medicine, 1954. ISSN 00222143. doi: 10.5555/uri:pii:0022214354901638.

[8] Alf S Alving, Charles F Johnson, Alvin R Tarlov, George J Brewer, Robert W Kellermeyer, and Paul E Carson. Mitigation of the haemolytic effect of primaquine and enhancement of its action against exoerythrocytic forms of the Chesson strain of Plasmodium vivax by intermittent regimens of drug administration: a preliminary report. Bulletin of the World Health Organization, 22(6):621, 1960.

[9] Sim Kheng, Sinoun Muth, Walter RJ Taylor, Narann Tops, Khem Kosal, Khon Sothea, Phum Souy, Saorin Kim, Chuor Meng Char, Chan Vanna, et al. Tolerability and safety of weekly primaquine against relapse of Plasmodium vivax in Cambodians with glucose-6-phosphate dehydrogenase deficiency. BMC Medicine, 13(1):1–10, 2015.

[10] Walter R J Taylor, Niamh Meagher, Benedikt Ley, Kamala Thriemer, Germana Bancone, Ari Satyagraha, Ashenafi Assefa, Krisin Chand, Nguyen Hoang Chau, Mehul Dhorda, Tamiru S Degaga, Lenny L Ekawati, Asrat Hailu, Mohammad Anwar Hasanzai, Mohammad Nader Naddim, Ayodhia Pitaloka Pasaribu, Awab Ghulam Rahim, Inge Sutanto, Ngo Viet Thanh, Nguyen Thi Tuyet-Trinh, Naomi Waithira, Adugna Woyessa, Arjen Dondorp, Lorenz von Seidlein, Julie A Simpson, Nicholas J White, J Kevin Baird, Nicholas P Day, and Ric N Price. Weekly primaquine for radical cure of patients with plasmodium vivax malaria and glucose-6-phosphate dehydrogenase deficiency. PLoS Neglected Tropical Diseases, 17(9):e0011522, September 2023.

[11] Sasithon Pukrittayakamee, Podjanee Jittamala, James A Watson, Borimas Hanboonkunupakarn, Pawanrat Leungsinsiri, Kittiyod Poovorawan, Kesinee Chotivanich, Germana Bancone, Cindy S Chu, Mallika Imwong, Nicholas PJ Day, Walter RJ Taylor, and Nicholas J White. Primaquine in glucose-6-phosphate dehydrogenase deficiency: an adaptive pharmacometric assessment of ascending dose regimens in healthy volunteers. eLife, 12:RP87318, feb 2024. ISSN 2050-084X. doi: 10.7554/eLife.87318. URL https://doi.org/10.7554/eLife.87318.

[12] James Watson, Walter RJ Taylor, Didier Menard, Sim Kheng, and Nicholas J White. Modelling primaquine-induced haemolysis in G6PD deficiency. eLife, 6:e23061, 2 2017. ISSN 2050-084X. doi: 10.7554/eLife.23061. URL https://doi.org/10.7554/eLife.23061.

[13] Angelo Minucci, Kamran Moradkhani, Ming Jing Hwang, Cecilia Zuppi, Bruno Giardina, and Ettore Capoluongo. Glucose-6-phosphate dehydrogenase (g6pd) mutations database: Review of the “old” and update of the new mutations. Blood Cells, Molecules, and Diseases, 48 (3):154–165, 2012. ISSN 1079-9796. doi: 10.1016/j.bcmd.2012.01.001. URL https://www.sciencedirect.com/science/article/pii/S1079979612000022.

[14] Robert S Franco. Measurement of red cell lifespan and aging. Transfus. Med. Hemother., 39 (5):302–307, October 2012.

[15] Robert J Commons, Megha Rajasekhar, Peta Edler, Tesfay Abreha, Ghulam R Awab, J Kevin Baird, Bridget E Barber, Cindy S Chu, Liwang Cui, André Daher Lilia Gonzalez-Ceron, Matthew J Grigg, Jimee Hwang, Harin Karunajeewa, Marcus V G Lacerda, Simone Ladeia-Andrade, Kartini Lidia, Alejandro Llanos-Cuentas, Rhea J Longley, Dhelio B Pereira, Ayodhia P Pasaribu, Sasithon Pukrittayakamee, Komal R Rijal, Inge Sutanto, Walter R J Taylor, Pham V Thanh, Kamala Thriemer, José Luiz F Vieira, James A Watson, Lina M Zuluaga-Idarraga, Nicholas J White, Philippe J Guerin, Julie A Simpson, Ric N Price, Bipin Adhikari, Nicholas M Anstey, Ashenafi Assefa, Sarah C Boyd, Nguyen Hoang Chau, Nicholas P J Day, Tamiru Shibiru Degaga, Arjen M Dondorp, Annette Erhart, Marcelo Urbano Ferreira, Prakash Ghimire, Justin A Green, Gavin Ckw Koh, Asrat Hailu Mekuria, Ivo Mueller, Mohammad Nader Naadim, Erni J Nelwan, Francois Nosten, David J Price, Jetsumon Sattabongkot, Kasia Stepniewska, Lorenz von Seidlein, Timothy William, Charles J Woodrow, and Adugna Woyessa. Effect of primaquine dose on the risk of recurrence in patients with uncomplicated plasmodium vivax: a systematic review and individual patient data meta-analysis. The Lancet Infectious Diseases, 24(2):172–183, February 2024.

[16] RS Hillman. Characteristics of marrow production and reticulocyte maturation in normal man in response to anemia. J. Clin. Invest., 48(3):443–453, March 1969.

[17] Nicholas J. White. Anaemia and malaria. Malaria Journal, 17(1):371, Oct 2018. ISSN 1475-2875. doi: 10.1186/s12936-018-2509-9. URL https://doi.org/10.1186/s12936-018-2509-9.

[18] James Watson, Walter R. J. Taylor, Germana Bancone, Cindy S. Chu, Podjanee Jittamala, and Nicholas J. White. Implications of current therapeutic restrictions for primaquine and tafenoquine in the radical cure of vivax malaria. PLOS Neglected Tropical Diseases, 12(4):1–14, 04 2018. doi: 10.1371/journal.pntd.0006440. URL https://doi.org/10.1371/journal.pntd.0006440.

[19] Elizabeth A Ashley, Judith Recht, and Nicholas J White. Primaquine: the risks and the benefits. Malaria Journal, 13(1):1–7, 2014.

[20] Judith Recht, Elizabeth A Ashley, and Nicholas J White. Use of primaquine and glucose-6-phosphate dehydrogenase deficiency testing: divergent policies and practices in malaria endemic countries. PLoS Neglected Tropical Diseases, 12(4):e0006230, 2018.

[21] Anatoly Kondrashin, Alla M Baranova, Elizabeth A Ashley, Judith Recht, Nicholas J White, and Vladimir P Sergiev. Mass primaquine treatment to eliminate vivax malaria: lessons from the past. Malar. J., 13:51, February 2014.

[22] Lucio Luzzatto, Mwashungi Ally, and Rosario Notaro. Glucose-6-phosphate dehydrogenase deficiency. Blood, 136(11):1225–1240, 09 2020. ISSN 0006-4971. doi: 10.1182/blood.2019000944. URL https://doi.org/10.1182/blood.2019000944.

[23] Inge Sutanto, Amin Soebandrio, Lenny L Ekawati, Krisin Chand, Rintis Noviyanti, Ari Winasti Satyagraha, Decy Subekti, Yulia Widya Santy, Chelzie Crenna-Darusallam, Instiaty Instiaty, Waras Budiman, Catur Bidik Prasetya, Soroy Lardo, Iqbal Elyazar, Stephan Duparc, Eve Cedar, Katie Rolfe, Disala Fernando, Alessandro Berni, Siôn Jones, Jörg-Peter Kleim, Kim Fletcher, Hema Sharma, Ana Martin, Maxine Taylor, Navin Goyal, Justin A Green, Lionel K Tan, and J Kevin Baird. Tafenoquine co-administered with dihydroartemisinin-piperaquine for the radical cure of plasmodium vivax malaria (INSPECTOR): a randomised, placebo-controlled, efficacy and safety study. Lancet Infect. Dis., 23 (10):1153–1163, October 2023.

[24] Parinaz Mehdipour, Megha Rajasekhar, Saber Dini, Sophie Zaloumis, Tesfay Abreha, Ishag Adam, Ghulam Rahim Awab, J Kevin Baird, Larissa W Brasil, Cindy S Chu, Liwang Cui, André Daher Margarete do Socorro M Gomes, Lilia Gonzalez-Ceron, Jimee Hwang, Harin Karunajeewa, Marcus V G Lacerda, Simone Ladeia-Andrade, Toby Leslie, Benedikt Ley, Kartini Lidia, Alejandro Llanos-Cuentas, Rhea J Longley, Wuelton Marcelo Monteiro, Dhelio B Pereira, Komal Raj Rijal, Kavitha Saravu, Inge Sutanto, Walter R J Taylor, Pham Vinh Thanh, Kamala Thriemer José, Luiz F Vieira, Nicholas J White, Lina M Zuluaga-Idarraga, Philippe J Guerin, Ric N Price, Julie A Simpson, Robert J Commons, and WWARN Vivax Adherence Study Group. Effect of adherence to primaquine on the risk of plasmodium vivax recurrence: a WorldWide antimalarial resistance network systematic review and individual patient data meta-analysis. Malar. J., 22(1):306, October 2023.

[25] Bob Carpenter, Andrew Gelman, Matthew D. Hoffman, Daniel Lee, Ben Goodrich, Michael Betancourt, Marcus Brubaker, Jiqiang Guo, Peter Li, and Allen Riddell. Stan: A probabilistic programming language. Journal of Statistical Software, 76(1):1–32, 2017. doi: 10.18637/jss.v076.i01.

[26] Daniel Lewandowski, Dorota Kurowicka, and Harry Joe. Generating random correlation matrices based on vines and extended onion method. Journal of Multivariate Analysis, 100(9):1989–2001, 2009. ISSN 0047-259X. doi: 10.1016/j.jmva.2009.04.008.

[27] R Core Team. R: A Language and Environment for Statistical Computing. R Foundation for Statistical Computing, Vienna, Austria, 2021. URL https://www.R-project.org/.

[28] Jonah Gabry, Rok Češnovar, and Andrew Johnson. cmdstanr: R Interface to ‘CmdStan’, 2024. https://mc-stan.org/cmdstanr/, https://discourse.mc-stan.org.

